# Development and Validation of a Multimodal Clinical, Pathologic, and Genomic Model for Breast Cancer Recurrence

**DOI:** 10.64898/2026.05.08.26352562

**Authors:** Ngoc-Kim Nguyen, Anran Li, Sara Kochanny, James Dolezal, Siddhi Ramesh, Gil Shamai, Junhan Zhao, Rita Nanda, Nan Chen, Olufunmilayo I. Olopade, Megan Sullivan, E Martin Flores, Galina Khramtsova, Sasha Jain-Liu, Riley Medenwald, Poornima Saha, Linda McCart, Mark Watson, W Fraser Symmans, Kevin Kalinsky, Lajos Pusztai, Michal Gala, Evan D. Paul, Barbora Huraiová, Pavol Čekan, Ann H. Partridge, Lisa Carey, Daniel Stover, Katharine Yao, Joseph A. Sparano, Dezheng Huo, Alexander T. Pearson, Frederick M. Howard

**Author notes:** **Corresponding Author**: Frederick Howard, MD, Assistant Professor, Section of Hematology Oncology, Department of Medicine, The University of Chicago, 5841 S Maryland Ave, Chicago, IL 60637.

## Abstract

**Purpose:** To develop and validate a multimodal recurrence-risk model integrating histology, genomic testing, and clinical variables.

**Methods:** We developed AI-Path, a whole-slide image biomarker for recurrence prediction trained in CALGB 9344, and validated it in three independent cohorts: TAILORx, a multi-site Chicago cohort, and the MDX-BRCA cohort. We then integrated AI-Path with Oncotype DX Recurrence Score (RS), tumor size, and nodal status into a Cox model, PathClinRS, fit using 60% of cases from TAILORx, with the remaining 40% held out for validation. The primary end point was distant recurrence-free interval. Performance was assessed using Harrell’s concordance index (C-index) and Kaplan-Meier analyses.

**Results:** A total of 12,418 patients were included. In TAILORx, AI-Path outperformed RS for distant recurrence (C-index, 0.682 vs 0.647; *P* = .038), driven by superior prediction of late recurrence (0.656 vs 0.567; *P* < .001). In node-negative disease, PathClinRS outperformed RSClin in the TAILORx fitting (0.72 vs 0.70; *P* = .016) and validation sets (0.74 vs 0.70; *P* = .004). In node-positive disease, PathClinRS outperformed RSClinN+ in Chicago (0.94 vs 0.74; *P* < .001) and MDX-BRCA (0.71 vs 0.66; *P* = .004) cohorts. Compared with NATALEE eligibility, PathClinRS identified nearly twice as many high-risk node-negative patients while maintaining a comparable 10-year distant recurrence risk (16.7% vs 16.6% per NATALEE eligibility in TAILORx fitting; 21.0% vs 19.4% in TAILORx validation). PathClinRS identified 68% of intermediate risk premenopausal patients as low-risk with no evidence of chemotherapy benefit, compared to only 36% identified as low risk by standard clinicopathologic criteria.

**Conclusion:** Digital histopathology provides prognostic information complementary to genomic assays and has the potential to personalize therapy beyond existing clinicogenomic tools.

## Introduction

Hormone receptor-positive, HER2-negative breast cancer is the most common molecular subtype of early breast cancer, and although overall outcomes are favorable, a subset of patients are at high risk for recurrence. In particular, patients with estrogen receptor-positive disease remain at risk for late relapse well beyond 5 years, creating an ongoing need for tools that better distinguish indolent tumors from those with persistent metastatic potential^1^.

Genomic assays have become central to treatment decision-making in this setting. The Oncotype DX 21-gene recurrence score (RS) is widely used to estimate recurrence risk and guide chemotherapy use in patients with early breast cancer. The TAILORx trial established that most women with node-negative disease and an RS of less than 26 derived little or no benefit from chemotherapy^2^, although there is ongoing debate about the benefit in higher-risk/node-positive premenopausal patients^3^. To improve risk estimation beyond genomics alone, clinicogenomic models such as RSClin^4^ and RSClinN+^5^ were developed by integrating recurrence score results with standard clinicopathologic variables. These models provide more individualized estimates of distant recurrence risk and chemotherapy benefit than either genomic or clinical factors alone, supporting their use as a more refined decision aid^6^.

Recent advances in computational pathology have shown that artificial intelligence applied to whole-slide images can capture grade, receptor status, and even molecular features such as gene expression patterns^7,8^. Numerous studies have suggested that routine hematoxylin and eosin (H&E) stained pathology slides capture robust prognostic information from tumor architecture, stromal context, differentiation, and immune infiltrate^9–14^. Although several models have achieved prognostic accuracy by directly predicting genomic risk scores from digital slides^10–12,15^, it is possible that there is additional prognostic information unique to digital pathology that would provide additive value to existing genomic testing.

At the same time, there is an increasing need for improved risk stratification and treatment-benefit prediction to better individualize the expanding array of adjuvant therapies. In addition to guiding chemotherapy decisions, better identification of high-risk patients may help refine selection for adjuvant treatment intensification strategies such as the use of CDK4/6 inhibitors^16–18^ or ovarian suppression^19^.

In this study, we developed and validated a whole-slide image-based artificial intelligence biomarker, AI-Path, for recurrence prediction in breast cancer, and integrated it with clinical and genomic variables into a unified score, PathClinRS. We hypothesized that histology-derived risk would provide prognostic information complementary to Oncotype DX RS and conventional clinical factors, with added value for identifying late recurrence risk and for improving selection of patients for treatment escalation or de-escalation.

## Methods

### Study Design and Cohorts

This retrospective study incorporated two clinical trials and two real-world cohorts (**Figure 1**). AI-Path model training was performed exclusively in Cancer and Leukemia Group B (CALGB) 9344 (AC ± paclitaxel, NCT00003519) including patients of all receptor subtypes, and a locked model was subsequently validated in TAILORx (NCT00310180). Two real-world cohorts were used for additional validation: a pooled cohort from three Chicago-area hospitals of patients diagnosed from 2010 - 2023 (including University of Chicago, Endeavor Health Cancer Institute and Ingalls Memorial Hospital), and the MultiplexDX breast cancer (MDX-BRCA) cohort^12,20^. Patients from the AI-Path validation cohorts were required to have invasive breast carcinoma, digitized H&E whole-slide images (WSI) from FFPE slides, Oncotype DX testing results (imputed score from gene expression used for MDX-BRCA as previously described^20^), and standard clinicopathologic covariates (age, tumor size, nodal status, recurrence status). Median imputation was performed for missing grade, and exact tumor size / nodal status was estimated from T/N stage in the MDX-BRCA cohort. The overall study design was approved by the University of Chicago institutional review board protocol IRB 22-0707; patients from the University of Chicago were prospectively consented on protocol IRB 16-352A.

**Figure 1.**
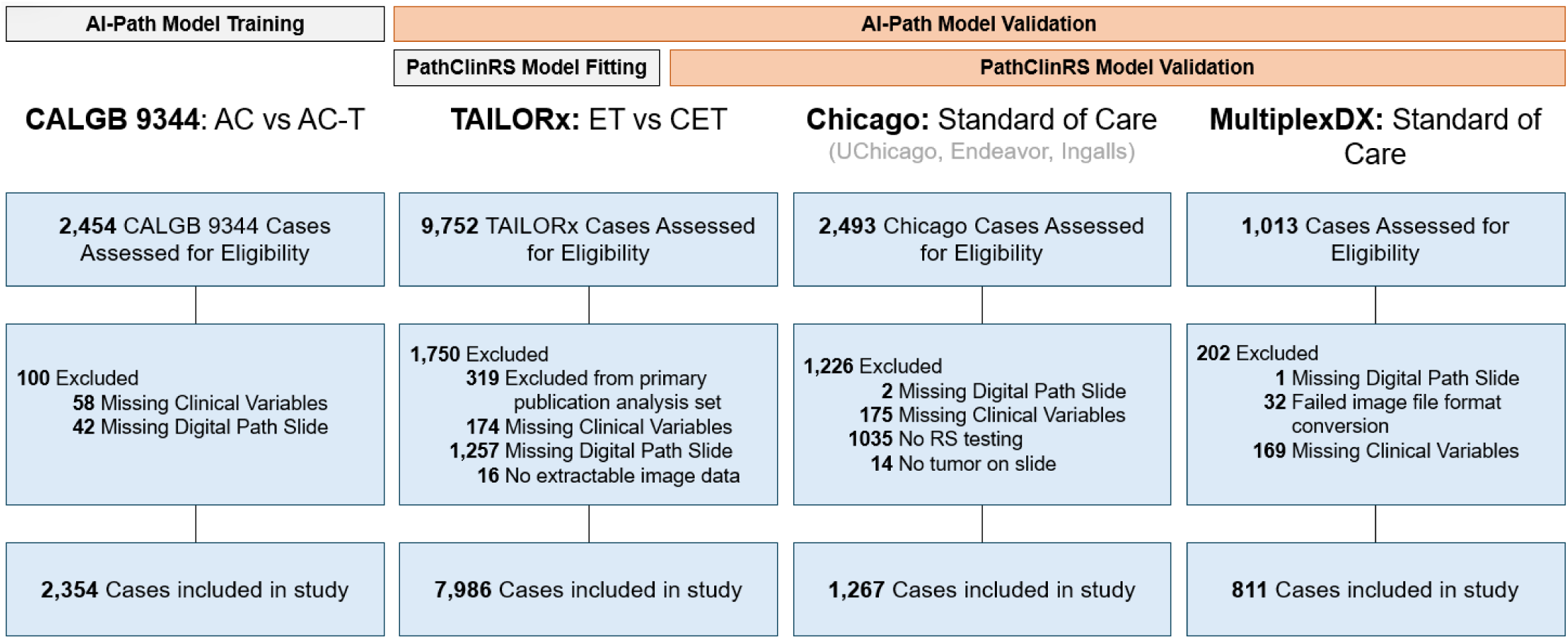
Consort Diagram of Cohorts Used for Model Training / Validation.

### Model Development

The AI-Path score was derived from digitized H&E whole-slide images using a survival framework implemented using the Slideflow pipeline with a Cox proportional hazards objective^21^. Whole-slide images were tiled into patches at an effective 10X resolution with Otsu-thresholding^22^ and Gaussian blur-based filtering^23^ for quality control. Tile-level feature representations were extracted with the H-optimus-0 foundation model^24^, and patient-level risk was learned using a transformer-based multiple-instance learning architecture^25^, after hyperparameter optimization with comparison to other foundation models^26,27^ (**Supplemental Table 1**). The resulting predictions are an unbounded estimate of relative log hazard, with 95% of results ranging from -2 to 2.

To determine whether AI-Path added prognostic information beyond RS and standard clinical variables, a multivariable Cox proportional hazards model, termed PathClinRS, was fit in the TAILORx model-fitting cohort constituting 60% of the dataset, with performance assessed in the held-out 40%. Candidate predictors included AI-Path, RS, age, tumor grade, estrogen receptor status, progesterone receptor status, and tumor size. Transforms of the input variables were selected to optimize the log likelihood statistic in univariable models fit in the model-fitting cohort. To extend the model to node-positive disease, the effect of positive lymph nodes was estimated using combined data from the TAILORx fitting cohort and CALGB 9344. Standard scalar normalization was applied to the MDX-BRCA cohort AI-Path outputs for multivariable model integration due to the use of a different slide scanner in this cohort. We also evaluated whether a genomic-free model could deliver comparable performance, utilizing the previously published model TIGER, trained on The Cancer Genome Atlas, that predicts Oncotype DX (as well as other gene expression signatures) from digital slide images^8,15^.

### Outcome Measures and Comparators

The primary end point was distant recurrence-free interval, defined as time from diagnosis or trial registration, according to source dataset convention, to distant recurrence, with censoring at last follow-up. Secondary end points included distant recurrence-free survival (DRFS) or overall survival (OS). Early recurrence was defined as recurrence within 5 years, and late recurrence as recurrence occurring after 5 years. The prognostic value of AI-Path was compared to Oncotype DX, and the prognostic value of PathClinRS was compared to RSClin risk estimates. For node-negative disease, the 10-year distant recurrence risk after aromatase inhibitor treatment from RSClin N0 was used to provide treatment-agnostic estimates. For node-positive disease, the RSClin N+ 5-year recurrence-free survival estimate for post-menopausal patients was used to allow for continuous estimates for all patients (as estimates are not given for high-risk premenopausal patients, and menopausal status was not collected in all cohorts). Number of positive nodes were capped at 3 for RSClin N+ calculations. For comparison to NATALEE eligibility, patients were classified as eligible with positive nodal status, tumor size greater than 5 cm, or tumor size greater than 2 cm with either grade 3 disease or RS at least 26. Ki-67 was not used due to lack of availability across datasets. For prediction of chemotherapy benefit in premenopausal patients, we compared performance to clinical-genomic risk as used in prior TAILORx analysis, with patients with RS 21-25 or RS 16-20 with high clinical risk classified as high risk.

### Statistical Analysis

Model discrimination for time-to-event outcomes was assessed using Harrell’s concordance index (C-index). For direct comparisons of models within the same cohort, paired bootstrap resampling was used to estimate 95% confidence intervals and p values for differences in C-index. Early / late recurrence was similarly quantified with C-index, and year-specific recurrence classification was summarized using time-dependent receiver operating characteristic analyses. To evaluate the joint prognostic effect of AI-Path and RS, logistic models with restricted cubic spline terms for both predictors were fit and used to visualize estimated recurrence risk across their joint distribution. Kaplan-Meier methods were used to estimate cumulative distant recurrence risk over time and at 10 years. For binary risk-group analyses, a cutoff derived from the TAILORx fitting cohort (85% for equivalence to NATALEE high risk criteria; 60% for premenopausal chemotherapy benefit) was applied unchanged to the held-out TAILORx validation cohort and to the external cohorts to classify patients as low or high risk by PathClinRS. Chemotherapy-benefit analyses were restricted to TAILORx patients randomized to endocrine therapy alone or chemoendocrine therapy. Distant recurrence risk was estimated within treatment-by-risk strata, and chemotherapy-associated hazard ratios and interaction p values were estimated from Cox models including treatment, risk group, and their interaction. All analyses were performed in Python using lifelines, scikit-learn, statsmodels, and Slideflow packages. Statistical tests were 2-sided, and p values less than .05 were considered statistically significant.

## Results

### Patient Demographics

A total of 12,418 patients were included across four cohorts (**Table 1**). TAILORx was entirely node-negative and hormone receptor positive/HER2 negative by design, whereas the Chicago and MDX-BRCA cohorts included both node-negative and node-positive cases. Mean RS was 18 in both TAILORx and Chicago cohorts and 26 in the MDX-BRCA cohort. Distant recurrence occurred in 5.8%, 2.3%, and 12.3% of patients in the TAILORx, Chicago, and MDX-BRCA cohorts; median follow-up was 10.3, 6.4, and 7.3 years respectively.

**Table 1.** Baseline Demographics of Included Cohorts.

|  |  | **CALGB9344 (n = 2354)** | **TAILORx (n = 7986)** | **Chicago (n = 1267)** | **MDX-BRCA (n = 811)** |
| --- | --- | --- | --- | --- | --- |
| **Age, mean (SD)** |  | 48.5 (9.8) | 55.6 (9.1) | 57.4 (9.9) | 59.4 (12.2) |
| **Tumor Size in mm, mean (SD)** |  | 31.4 (18.9) | 17.5 (8.3) | 19.8 (12.4) | 23.2 (14.5) |
| **Involved Nodes, n (%)** | 0 | 1 (0.0) | 7986 (100.0) | 1080 (85.2) | 483 (59.6) |
|  | 1 – 3 | 1114 (47.3) |  | 185 (14.6) | 254 (31.3) |
|  | 4 – 9 | 969 (41.2) |  | 2 (0.2) | 45 (5.5) |
|  | 10+ | 270 (11.5) |  |  | 29 (3.6) |
| **HR, n (%)** | Positive | 1584 (67.3) | 7986 (100.0) | 1256 (99.1) | 604 (74.5) |
|  | Negative | 770 (32.7) |  | 11 (0.9) | 207 (25.5) |
| **HER2, n (%)** | Positive |  |  | 9 (0.7) | 135 (16.6) |
|  | Negative |  | 7986 (100.0) | 896 (70.7) | 676 (83.4) |
|  | Unknown | 2354 (100.0) |  | 362 (28.6) |  |
| **Grade, n (%)** | 1 |  | 2083 (26.1) | 269 (21.2) | 112 (13.8) |
|  | 2 |  | 4318 (54.1) | 662 (52.2) | 348 (42.9) |
|  | 3 |  | 1384 (17.3) | 206 (16.3) | 343 (42.3) |
|  | Unknown | 2354 (100.0) | 201 (2.5) | 130 (10.3) | 8 (1.0) |
| **RS, n (%)** | 0 – 10 |  | 1337 (16.7) | 263 (20.8) | 226 (27.9) |
|  | 11 – 25 |  | 5490 (68.7) | 785 (62.0) | 194 (23.9) |
|  | 26 – 100 |  | 1159 (14.5) | 219 (17.3) | 391 (48.2) |
|  | Unknown | 2354 (100.0) |  |  |  |
| **Distant Recurrence, n (%)** | No | 1495 (63.5) | 7520 (94.2) | 1238 (97.7) | 711 (87.7) |
|  | Yes | 859 (36.5) | 466 (5.8) | 29 (2.3) | 100 (12.3) |

### Prognostic Patterns of the AI-Path Model

We first compared discrimination for distant recurrence using AI-Path and Oncotype DX across cohorts (**Figure 2 A-C, Supplemental Table 2**). In TAILORx, AI-Path showed better overall discrimination than Oncotype DX (C-index, 0.682 vs 0.647, *P* = .038); although this was driven by improvements in prediction of late recurrence (C-index 0.656 vs 0.567, *P* < .001). Consistently higher prognostic accuracy in TAILORx was seen across subgroups (**Supplemental Figure 1**), including those with low RS (≤ 10, AI-Path C-index 0.615 vs RS C-index 0.549), intermediate RS (11-25, C-index 0.640 vs 0.571) and high RS (>26, C-index 0.630 vs 0.607). AI-Path also outperformed Oncotype DX for prediction of late recurrence in the MDX-BRCA cohort (C-index 0.597 vs 0.464, *P* < 0.001). Similar patterns were seen in the Chicago cohort although no comparison was statistically significant given smaller number of events and shorter follow-up; and similar findings were seen across cohorts for associations with DRFS and OS (**Supplemental Figure 2**). Year-specific AUC for distant recurrence in TAILORx demonstrates the prognostic value of AI-Path exceeds that of Oncotype for all years after year 5 (**Figure 2D**). Despite the similar predictive accuracy for early recurrence, AI-Path is poorly correlated with Oncotype DX (Pearson *r* = 0.29), suggesting that the two tests provide independent value (**Figure 2E**). Spline estimates of distant recurrence rates suggest that AI-Path and Oncotype have additive value for early recurrence, whereas late recurrence is predominantly determined by AI-Path (**Figure 2F-H**).

**Figure 2.**
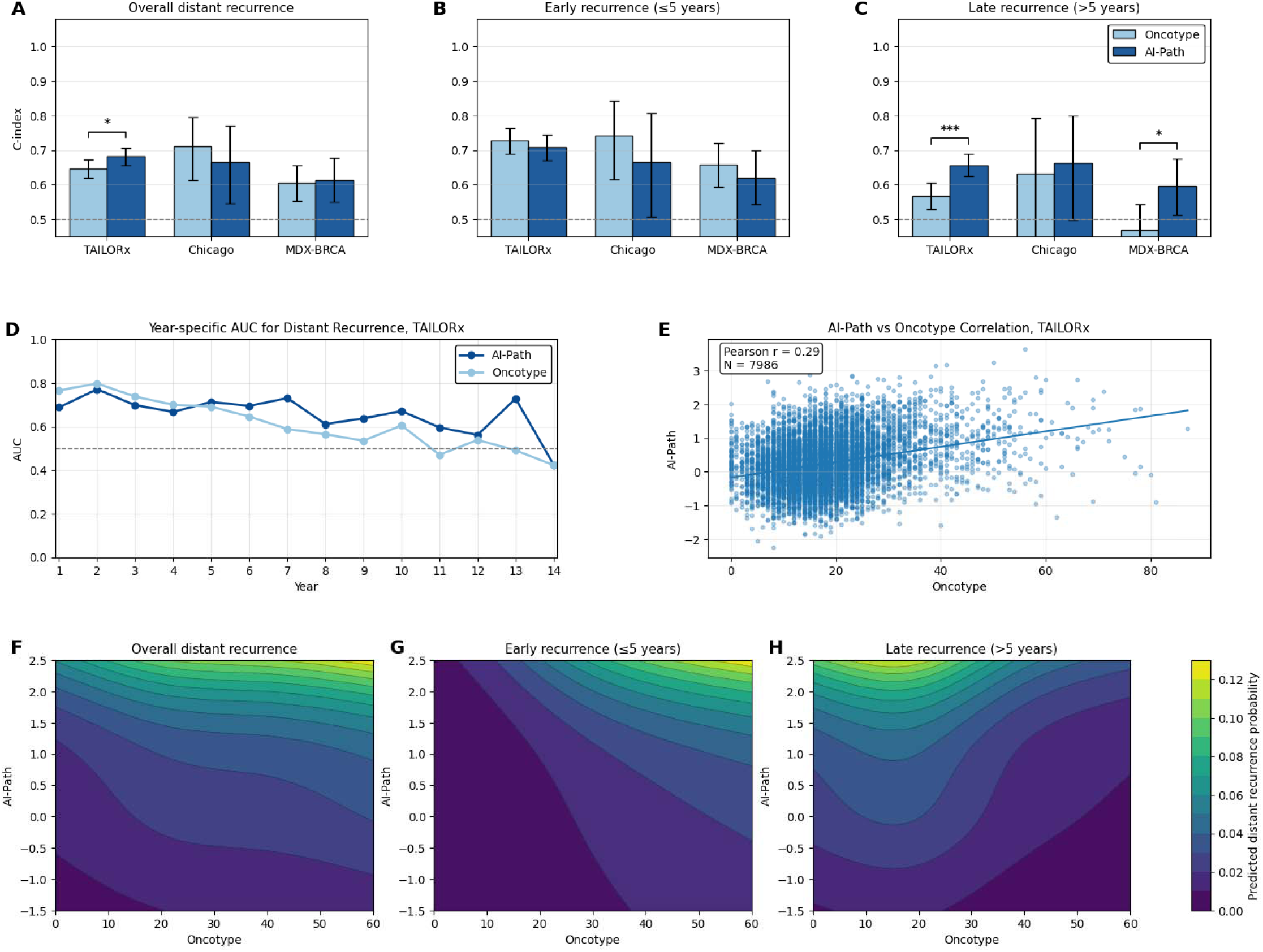
AI-Path and Oncotype show Distinct and Complementary Associations with Recurrence. (**A–C**) C-index estimates for AI-Path and Oncotype across the TAILORx, Chicago, and MDX-BRCA datasets for overall, early, and late distant recurrence. (**D**) Year-specific AUC for distant recurrence in TAILORx. (**E**) Correlation between AI-Path and Oncotype scores in TAILORx. (**F-H**) Restricted cubic spline surfaces depicting predicted distant recurrence probability across the joint distribution of Oncotype and AI-Path scores for overall, early, and late recurrence. * P < 0.05; ** P < 0.01; *** P < 0.001

To understand the factors contributing to AI-Path’s predictions, we first evaluated heatmaps of model attention and model prediction values for predicted high / low risk cases (**Figure 3A-B**). Risk predictions were associated with poor differentiation with increased mitoses and nuclear pleomorphism, and lower immune infiltrate. Furthermore, we leveraged HistoXGAN^28^, an approach to produce synthetic images corresponding to low-risk or high-risk model predictions from a set of base images (**Figure 3C**) – demonstrating that in addition to high grade features, risk was associated with lymphovascular invasion, reduced lymphocytic infiltrate, and increased tumor vascularization.

**Figure 3.**
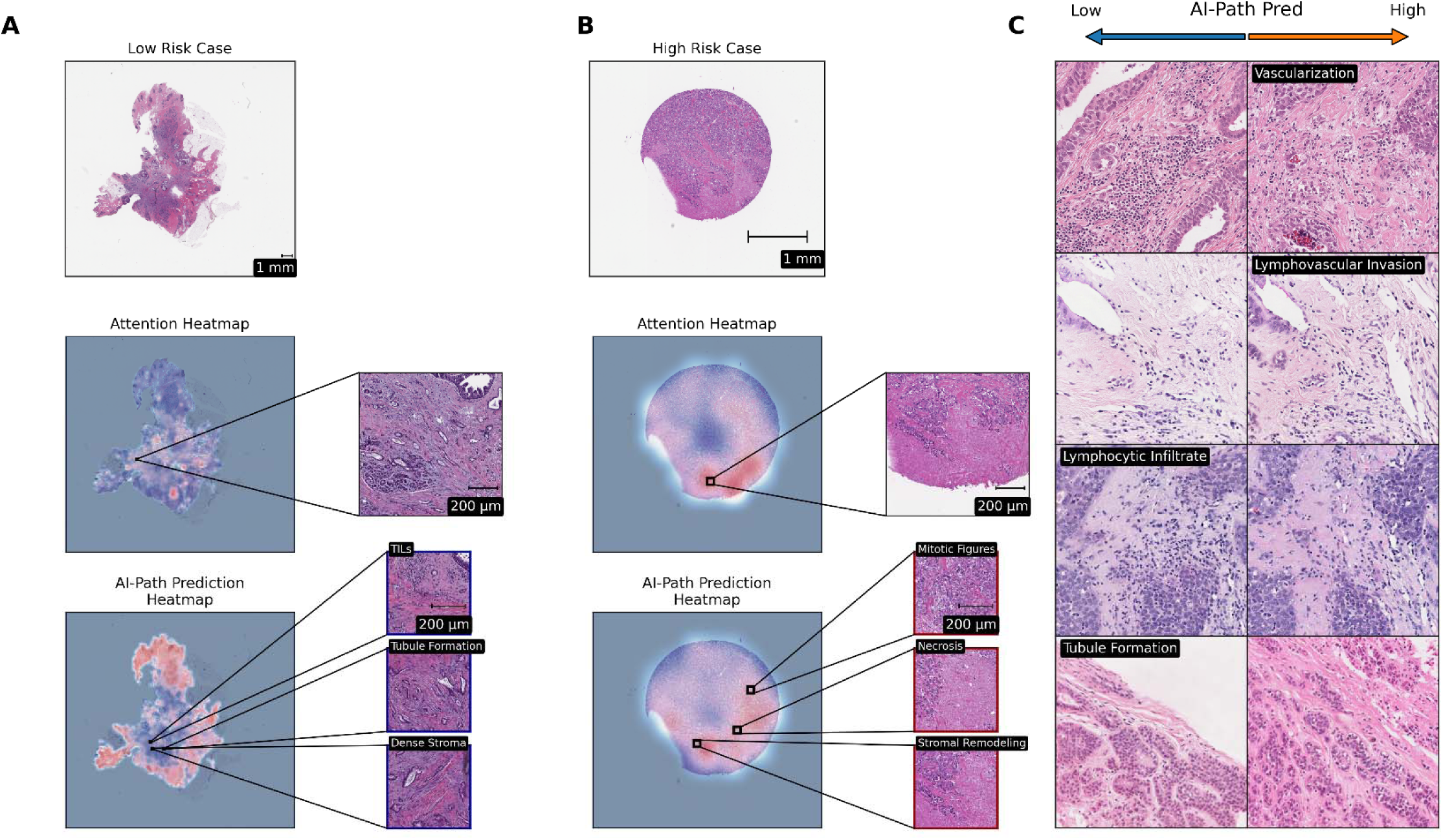
Model Explainability Analysis. Shown are model predictions for a representative (**A**) low-risk case and (**B**) high-risk case. An attention heatmap illustrates slide areas that contribute to model predictions (with red indicating higher attention). The prediction heatmap (with red highlighting a prediction of higher risk by AI-Path), illustrates the model signature prediction for each region of the slide. (**C**) Digital histology images were generated from representative cases and subsequently modified in a stepwise manner using the HistoXGAN pipeline until AI-Path model predictions reached low or high risk levels. For each pair, the images on the left and right represent low- and high-risk versions of the same baseline image, respectively, with key features annotated.

### A Unified Clinical, Pathologic, and Genomic Model for Recurrence Risk

In the TAILORx model fitting cohort, multivariable modeling showed that AI-Path, RS, and tumor size each remained independently associated with recurrence risk, whereas age, grade, and receptor status did not (**Figure 4A, Supplemental Table 3**). We thus fit a three-parameter Cox model in the TAILORx model fitting cohort - with hazard ratios (per SD) of 1.50 (95% CI, 1.33 to 1.69) for AI-Path, 1.35 (95% CI, 1.21 to 1.51) for RS, and 1.41 (95% CI, 1.26 to 1.57) for tumor size. The coefficient for number of involved nodes was estimated from TAILORx fitting and CALGB 9344 cohorts and scaled for inclusion in a final pathologic, clinical, and genomic model (PathClinRS) using the relative tumor size hazard ratios in each dataset (hazard ratio per SD, 1.91; 95% CI, 1.52 to 2.41; *P* < .001).

**Figure 4.**
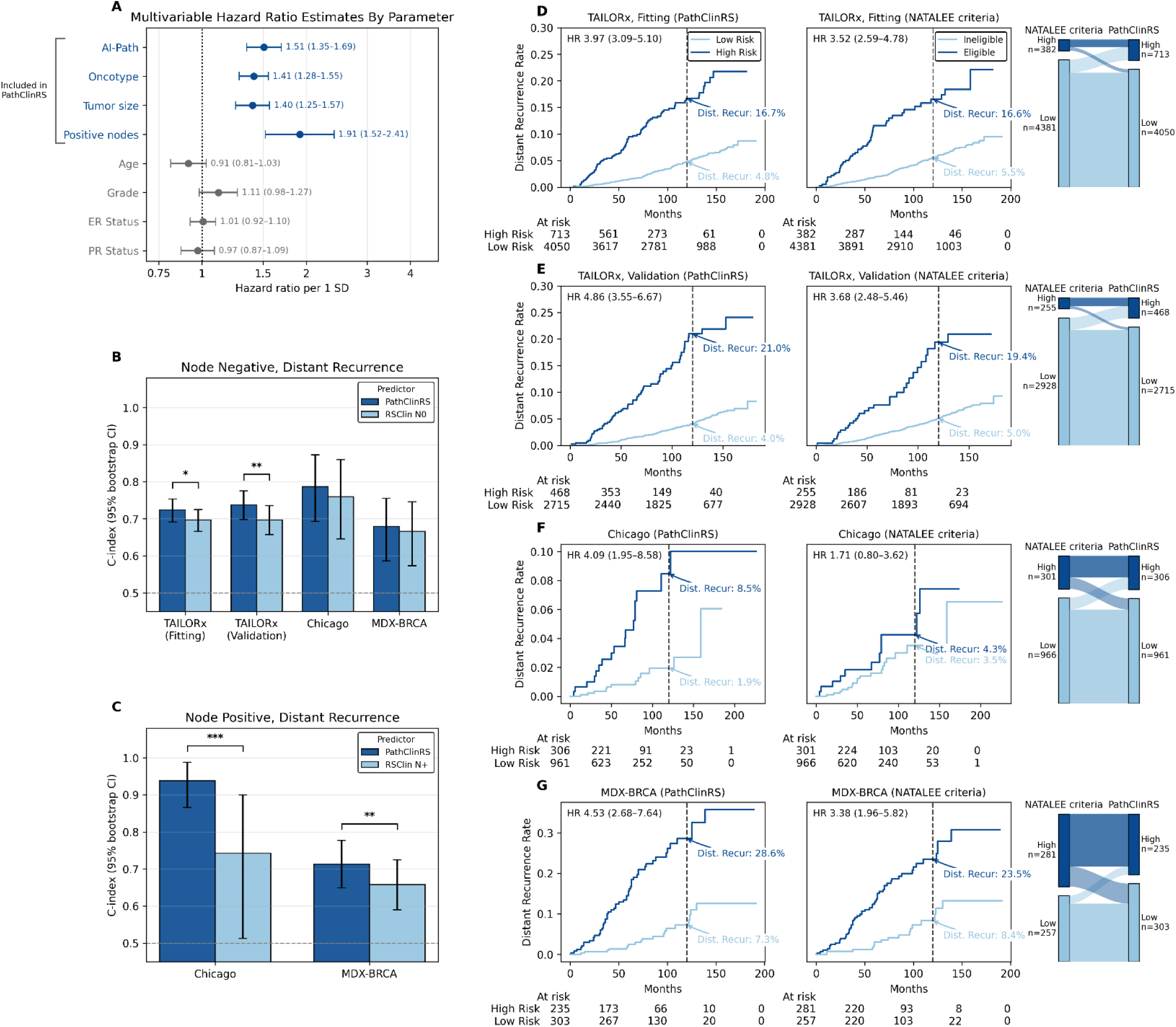
PathClinRS improves Prognostic Discrimination and Risk Stratification. **(A)** Multivariable hazard ratio (HR) estimates for distant recurrence risk per 1 standard deviation (SD) increase in each parameter in the TAILORx Cox model fitting cohort. AI-Path, Oncotype, tumor size, and positive nodes were retained in the PathClinRS model. The coefficient for number of involved nodes was estimated using relative hazard of tumor size and number of involved nodes in a Cox model from the TAILORx fitting cohort and CALGB 9344. (**B-C**) Predictive performance for overall distant recurrence measured by C-index in node-negative cohorts (**B**) and node-positive cohorts (**C**), comparing PathClinRS with RSClin N0 and RSClin N+. (**D-G**) Kaplan-Meier estimates of distant recurrence in low- and high-risk groups defined by PathClinRS (left) or NATALEE trial eligibility criteria (right) in the TAILORx model fitting cohort (**D**), TAILORx validation cohort (**E**), Chicago cohort (**F**), and MDX-BRCA cohort (**G**). HRs for high- versus low-risk groups are shown in each panel, with corresponding distant recurrence rates annotated on the curves. Sankey plots showing the reclassification of cases per NATALEE eligibility versus PathClinRS high-risk are shown. Across cohorts, PathClinRS showed improved or comparable discrimination relative to clinical comparator models and provided clear separation of low- and high-risk groups. * P < 0.05; ** P < 0.01; *** P < 0.001

In node-negative cohorts, PathClinRS significantly outperformed RSClin in both the TAILORx fitting cohort (C-index, 0.72 vs 0.70, *P* = .016) and the TAILORx validation cohort (0.74 vs 0.70; *P* = .004). Numerically higher discriminative value was seen in the Chicago (0.79 vs 0.76, *P* = 0.404) and MDX-BRCA (0.68 vs 0.67, *P* = 0.502) cohorts, although these differences were not statistically significant (**Figure 4B, Supplemental Table 4**). In node-positive cohorts, PathClinRS similarly demonstrated consistently superior performance, in both the Chicago (0.94 vs 0.74, *P* < 0.001) and MDX-BRCA (0.71 vs 0.66, *P* = 0.004) cohorts (**Figure 4C**). Similar findings were seen with respect to DRFS and OS (**Supplemental Figure 3**).

Although most patients with hormone receptor positive breast cancer undergo genomic testing, there are scenarios where such testing is unavailable. Thus, we also evaluated a previously developed model for Oncotype DX (as well as other gene expression signatures), as a surrogate for RS in PathClinRS^8^. This histologic model achieved an AUROC of 0.85 for high RS (≥ 26) in the TAILORx cohort (**Supplemental Figure 4A**) and had independent prognostic value from AI-Path (**Supplemental Figure 4B**). AI-RS (C-index 0.661) had similar accuracy for distant recurrence as RS (C-index 0.647) in TAILORx. Performance of PathClinRS, substituting the pathology-based Oncotype prediction for genomic RS, was similar (although slightly numerically lower in most cases) to the model with genomic Oncotype values (**Supplemental Figure 4C-D**).

### Identification of High-Risk Patients for Treatment Intensification

We evaluated model cutoffs in the TAILORx fitting cohort that would select patients with similar distant recurrence risk as the NATALEE eligibility criteria to potentially improve selection of patients who may benefit from treatment intensification with CDK4/6 inhibitors (**Supplemental Figure 5**). We found the 85^th^ percentile identified the largest proportion of patients that had a similarly elevated risk as NATALEE-eligible patients (**Supplemental Figure 5**). This cutoff was then applied to HR+/HER2- patients from external cohorts (**Figure 4 D-G, Supplemental Table 5**). This cutoff identified a high-risk population with a 16.7%, 21.0%, 8.5%, and 28.6% risk of distant recurrence in the TAILORx fitting, TAILORx validation, Chicago, and MDX-BRCA cohorts; indicating similar or higher risk to patients eligible for NATALEE in these cohorts (16.6%, 19.4%, 4.3%, 23.5% respectively). This cutoff identified approximately twice as many patients as ‘high risk’ (as compared to the NATALEE criteria) in TAILORx with similarly high distant recurrence risks. Conversely, the total number of ‘high risk’ patients was similar in the Chicago / MDX-BRCA cohorts – in these cohorts, increases in high-risk node-negative patients were balanced by decreases in high-risk node-positive patients. These cutoffs were also more discriminative of risk (HR 3.97, 4.86, 4.09, and 4.53 in fitting / validation / Chicago / MDX-BRCA) than NATALEE eligibility (HR 3.52, 3.68, 1.71, and 3.38). Consistently high risk was seen among the subset of patients classified as high risk by PathClinRS that would be ineligible for NATALEE, as well as lower risk among those eligible for NATALEE but low risk by PathClinRS (**Supplemental Figure 6, Supplemental Table 6**).

### Avoiding Overtreatment in Premenopausal Patients

We also evaluated whether PathClinRS could be used to de-escalate treatment for premenopausal women who may be recommended for treatment escalation per standard clinicogenomic risk criteria. We identified the lowest proportion of premenopausal patients for which no chemotherapy benefit was seen in the TAILORx model fitting cohort, demonstrating that a chemotherapy benefit begins to emerge with PathClinRS scores above the 60^th^ percentile (**Supplemental Figure 7**). Thus, we used this percentile defined by the model fitting cohort to stratify patients into low / high chemotherapy benefit groups. Traditional clinical-genomic high-risk criteria identified ∼64% of the intermediate risk (TAILORx arm B / C) patients as high risk, versus 32% (624) via the 60^th^ percentile cutoff of PathClinRS. Distant recurrence rates at 10 years remain low in the clinical-genomic low-risk group (2.2% with chemotherapy, 1.6% without); as well as in the PathClinRS low risk patients (2.7% with chemotherapy, 2.2% without in model fitting cohort; 1.9% with chemotherapy, 2.4% without in the validation cohort, **Figure 5, Supplemental Table 7**). Conversely, there was a 3.7% improvement in 10-year distant recurrence with chemotherapy for high-risk patients by clinical-genomic criteria, whereas the improvement for high-risk patients with PathClinRS was 6.9% in the model fitting and 6% in the validation cohorts. There was a trend towards interaction with PathClinRS high / low (P = 0.051) in the fitting cohort, but not in the validation cohort (P = 0.489). Applying this cutoff to postmenopausal patients was prognostic but did not identify a group with chemotherapy benefit (**Supplemental Figure 7**, **Supplemental Table 8**).

**Figure 5.**
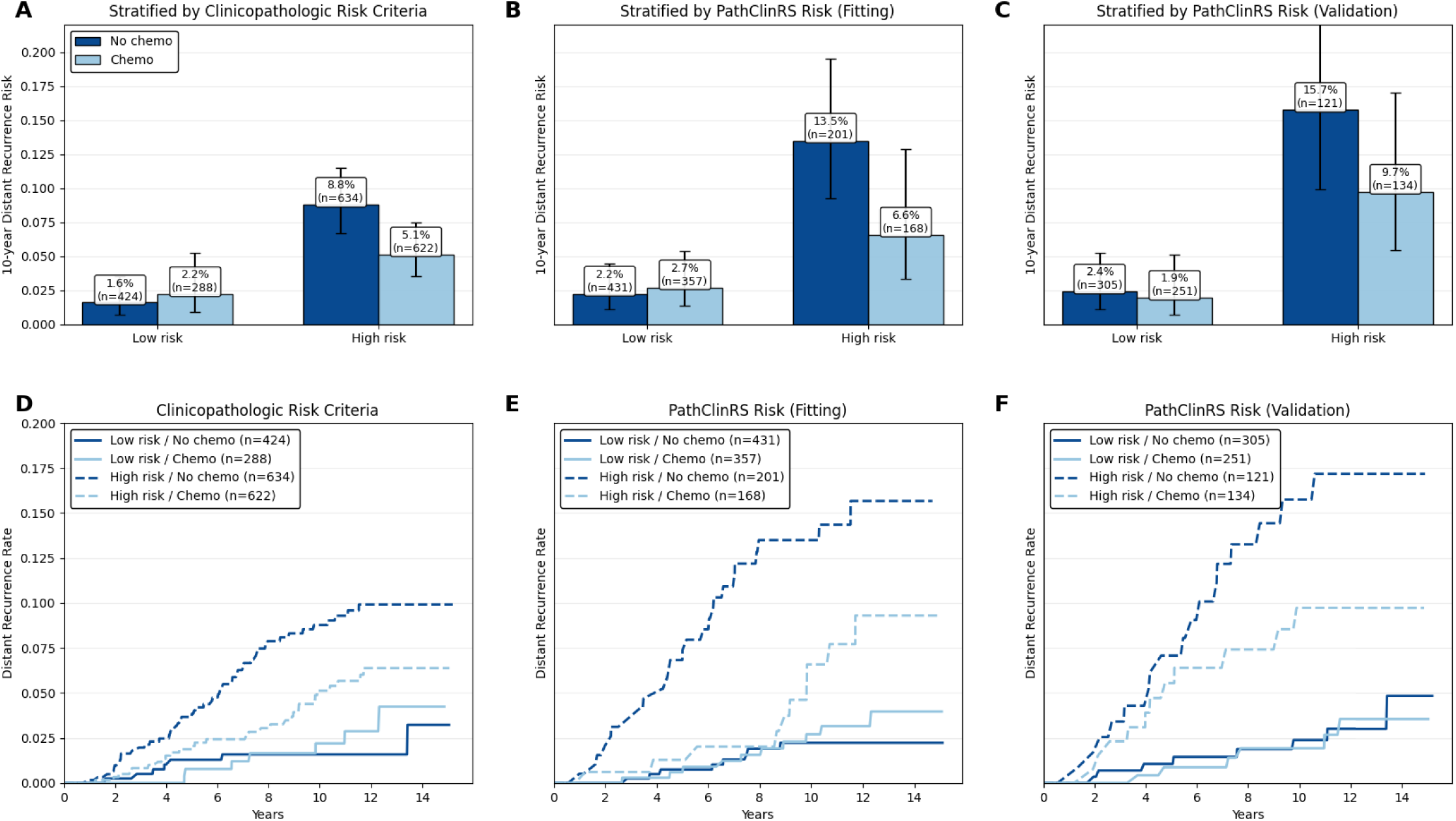
PathClinRS Predicts Chemotherapy Benefit in Intermediate Risk Premenopausal Patients in TAILORx. (**A-C**) Bar plots showing Kaplan-Meier-estimated 10-year distant recurrence risk in low- and high-risk premenopausal patients, further stratified by chemotherapy receipt. (**A**) Risk classification by clinicopathologic risk criteria, where high risk is defined as RS 21-25 or RS 16-20 with high clinical risk. (**B/C**) Risk classification by PathClinRS. (**D-F**) Kaplan-Meier curves for distant recurrence over time for the corresponding risk groups and chemotherapy strata for (**D**) clinicopathologic risk criteria and (**E/F**) PathClinRS risk groups.

## Discussion

In this multi-cohort study, a whole-slide image biomarker derived from routine H&E histology provided prognostic information that was complementary to the Oncotype DX recurrence score, with the most pronounced advantage in predicting late recurrence. AI-Path was only modestly correlated with Oncotype DX in TAILORx (r = 0.29), yet improved overall discrimination relative to Oncotype DX, largely because of stronger performance after year 5. When integrated with recurrence score and clinicopathologic factors, PathClinRS demonstrated consistently higher overall prognostic accuracy than RSClin risk predictions. More accurate risk prediction – especially prediction of late recurrence – may allow for more accurate personalization of treatments including extended endocrine therapy, adjuvant CDK4/6 inhibitors, and use of chemotherapy or ovarian suppression in premenopausal women.

These findings add to a rapidly emerging literature showing that computational pathology can refine risk assessment in HR-positive, HER2-negative early breast cancer. Although digital pathology can achieve equivalent prognostic accuracy to Oncotype DX^10^, we demonstrate here that there is additional prognostic information within digital slide images beyond what is captured by traditional genomic risk assays. Several recent models have demonstrated that digital pathology can improve the prediction of distant recurrence risk. In some cases, there is suggestion that multimodal models, incorporating digital pathology and clinical risk factors can outperform Oncotype DX. Our data extend that observation by benchmarking our AI-Path tumor biology model directly against Oncotype DX, without external clinical factors, and demonstrating superior long-term prognostic accuracy in the large randomized TAILORx cohort. Additionally, we demonstrate superior performance to existing genomic + clinical risk tools such as RSClin and RSClin N+, which may allow for further refined risk estimation.

In a recent TAILORx-based study, a multimodal model integrated image, clinical, and an expanded molecular panel derived from commercial assays including EndoPredict, PAM50, Breast Cancer Index, and MammaPrint^29^. This model achieved stronger prognostic performance than Oncotype DX for both overall and late recurrence in training and holdout validation sets, with a C-index of 0.71 in training and 0.73 on validation. Here, we demonstrate comparable / improved performance (C-index 0.72 on training, 0.74 on validation) without the use of additional molecular profiling which would be impractical in standard practice. Additionally, our image-only AI-Path model (C-index 0.682) outperformed the image-only model from this prior TAILORx study (C-index 0.632). Furthermore, we found pathology models separately conditioned on overall recurrence (AI-Path) and Oncotype (AI-RS) may perform similarly to our full multimodal model without the need for any genomic testing – which can be valuable for use in low-resource settings.

There are several important findings that may lay the groundwork for future escalation or de-escalation studies. First, PathClinRS identified groups with distant recurrence risk comparable to or greater than NATALEE-eligible populations, and refined risk estimates by identifying more node-negative and fewer node-positive patients. There has been extensive debate regarding whether CDK4/6 inhibitors are appropriate for high-risk node-negative patients^30^. To date, there are no clear biomarkers to predict relative benefit from CDK4/6 inhibitors in the adjuvant setting^31^ - but across trials and subgroups, adjuvant CDK4/6 inhibitors improve invasive disease-free survival by approximately 30%. Although ctDNA can perhaps identify the patients at highest risk, only a small proportion of patients with recurrence have baseline positivity with current-generation assays^32^. Thus, more accurate estimation of baseline risk (and therefore treatment benefit), can allow more personalized decision making regarding this treatment.

Furthermore, among premenopausal patients with intermediate recurrence scores in TAILORx, PathClinRS selected a smaller high-risk subset than conventional clinical-genomic criteria, while enriching for a larger absolute difference in 10-year distant recurrence with chemotherapy. Together, these findings suggest a potential role for pathology-based AI both in treatment intensification and in avoidance of overtreatment.

This study is closely aligned with the guidelines for studies testing AI models, as PathClinRS is positioned to guide treatment decisions, was validated externally beyond the development set, and was benchmarked against established clinical comparators^33^. Its main remaining gap relative to an implementation-focused standard is that clinical utility has not yet been tested prospectively. The external validation cohorts were small, with different follow-up horizons that may limit definite conclusions about the true external performance of PathClinRS. Additionally, the PathClinRS model was fit in a subset of TAILORx data - but as only three parameters (hazard ratios for the AI-Path, RS, and tumor size) were fit in this cohort, we believe there is minimal opportunity for overfitting.

Overall, these results support a pragmatic model for improving recurrence-risk assessment in HR-positive, HER2-negative early breast cancer. Compared with other recent multimodal approaches, we demonstrate that digital pathology adds independent and perhaps superior prognostic value to genomic assessment of tumor risk. With further validation, this approach could provide a scalable means to improve personalization of chemotherapy, endocrine therapy duration, and adjuvant treatment escalation.

## Data Availability

Data from CALGB 9344 are available upon approval of a data-sharing request submitted to the Alliance for Clinical Trials in Oncology via. The TAILORx dataset is available upon request from the ECOG-ACRIN Cancer Research Group or through the NCTN/NCORP Data Archive. RNA-sequencing data and a de-identified clinicopathological data table from the retrospective MDX-BRCA cohort have been deposited in the Gene Expression Omnibus (GEO) database under accession code GSE283522. H&E images from the MDX-BRCA dataset are available to qualified researchers following an initial request to. Extracted image features and clinical data from the Chicago cohort are available at https://doi.org/10.5281/zenodo.19956761, with additional data available from study authors upon reasonable request.

## Code Availability

The pipeline used for model training is available at https://github.com/fmhoward/slideflow-survival. The digital pathology model used in the analysis of PathClinRS with pathology estimates of Oncotype is available at https://github.com/fmhoward/TIGER.

## Acknowledgements

Data from Cancer and Leukemia Group B (CALGB) study 9344 were obtained from the Alliance for Clinical Trials in Oncology, a National Clinical Trials Network cooperative group, under Alliance data sharing agreement A152324; CALGB 9344 was supported in part by the National Cancer Institute of the National Institutes of Health (NIH) under Award Numbers U10CA180821 and U10CA180882 (to the Alliance for Clinical Trials in Oncology). TAILORx was conducted by the ECOG-ACRIN Cancer Research Group (Peter J. O’Dwyer, MD and Mitchell D. Schnall, MD, PhD, Group Co-Chairs) and supported by the National Cancer Institute of the National Institutes of Health under award numbers: U10CA180820, UG1CA233327, UG1CA189847, UG1CA233339, UG1CA233331, UG1CA233329, UG1CA233180 and UG1CA233320. The MDX-BRCA cohort from MultiplexDX was partially funded by European Union’s Horizon 2020 research and innovation program under an EIC Accelerator grant (946693, PČ) and the EU NextGenerationEU through the Recovery and Resilience Plan for Slovakia (09I01-03-V01-00003, PČ and 09I03-03-V03-00101, EDP). The content is solely the responsibility of the authors and does not necessarily represent the official views of the National Institutes of Health.

This work was also supported by the following research grants:

National Cancer Institute grant K08CA283261 (FMH)

Lynn Sage Breast Cancer Foundation (FMH)

Alliance Foundation Trials Special Projects Award (FMH)

Breast Cancer Research Foundation Next Generation Award (FMH)

National Institute of Dental and Craniofacial Research grant R56DE030958 (ATP)

National Cancer Institute grant R01CA276652 (ATP)

Department of Defense grant BC211095P1 (FMH, ATP, DH)

## Authors’ Disclosures of Potential Conflicts of Interest

JD reports ownership interest in Slideflow Labs. SR reports ownership interest in Slideflow Labs. NC reports consultant fees from Novartis, Guardant Health, Seagen, Stemline, AstraZeneca, and Daiichi Sankyo; and research funding from Eli Lilly, Olema, and Puma. JZ reports an advisory role and receipt of consulting fees and equity from Encapsulate Inc., equity in Alvus Health Inc., and serves on the research advisory board of Shriners Children’s. RN reports consulting funding from Astrazeneca, Daiichi Sankyo, Exact Sciences, GE, Gilead, Guardant Health, Merck, Moderna, Novartis, OBI, Pfizer, Sanofi, Seagen, Stemline, Summit Therapeutics and research funding from Arvinas, AstraZeneca, BMS, Corcept Therapeutics, Genentech/Roche, Gilead, GSK, Merck, Novartis, OBI Pharma, OncoSec, Pfizer, Relay, Seagen, Sun Pharma, Taiho. WFS reports stock / ownership interests in ISIS Pharmaceuticals, Delphi Diagnostics, and Eiger BioPharmaceuticals, consulting / advisory role for AstraZeneca, SAGA Diagnostics, and other uncompensated relationships with Delphi Diagnostics. KK reports consulting/advisory/speaker role for Daiichi Sankyo, Eli Lilly, Pfizer, Novartis, Eisai, AstraZeneca, Immunomedics, Merck, Seattle Genetics, Cyclacel, OncoSec, 4D Pharma, Puma, Genentech, Ascendant, Myovant, Takeda, and Menarini; and owns Grail stock options. LP reports has received consulting fees and honoraria for advisory board participation from Pfizer, Astra Zeneca, Merck, Novartis, Bristol-Myers Squibb, Stemline-Menarini, GlaxoSmithKline, Genentech/Roche, Personalis, Daiichi, Natera, Agendia, Exact Sciences, Radionetics, and institutional research funding from Seagen, GlaxoSmithKline, AstraZeneca, Merck, Pfizer and Bristol Myers Squibb. DS reports consulting / advisory role for Guardant Health and Natera, research funding from NeoGenomics Laboratories and Foundation Medicine. LC reports research funding from NanoString Technologies, Seagen, Veracyte, Gilead Sciences, Novartis, and other uncompensated relationships with Lilly, SeaGen, Gilead Sciences, Reveal Genomics, Novartis. OIO reports support from Healthy Life for All Foundation, other support from CancerIQ, grants and other support from Tempus, and grants from Color Genomics outside the submitted work. JS has received consulting fees from Genomic Health. ATP reports consulting fees from Prelude Biotherapeutics, LLC, Ayala Pharmaceuticals, Elvar Therapeutics, Abbvie, and Privo, and contracted research with Kura Oncology, Abbvie, and EMD Serono. FMH reports consulting fees from Novartis and Leica Biosystems, advisory board participation with Exact Sciences, and stock ownership with Celcuity.

---

**Supplemental Figure 1.**
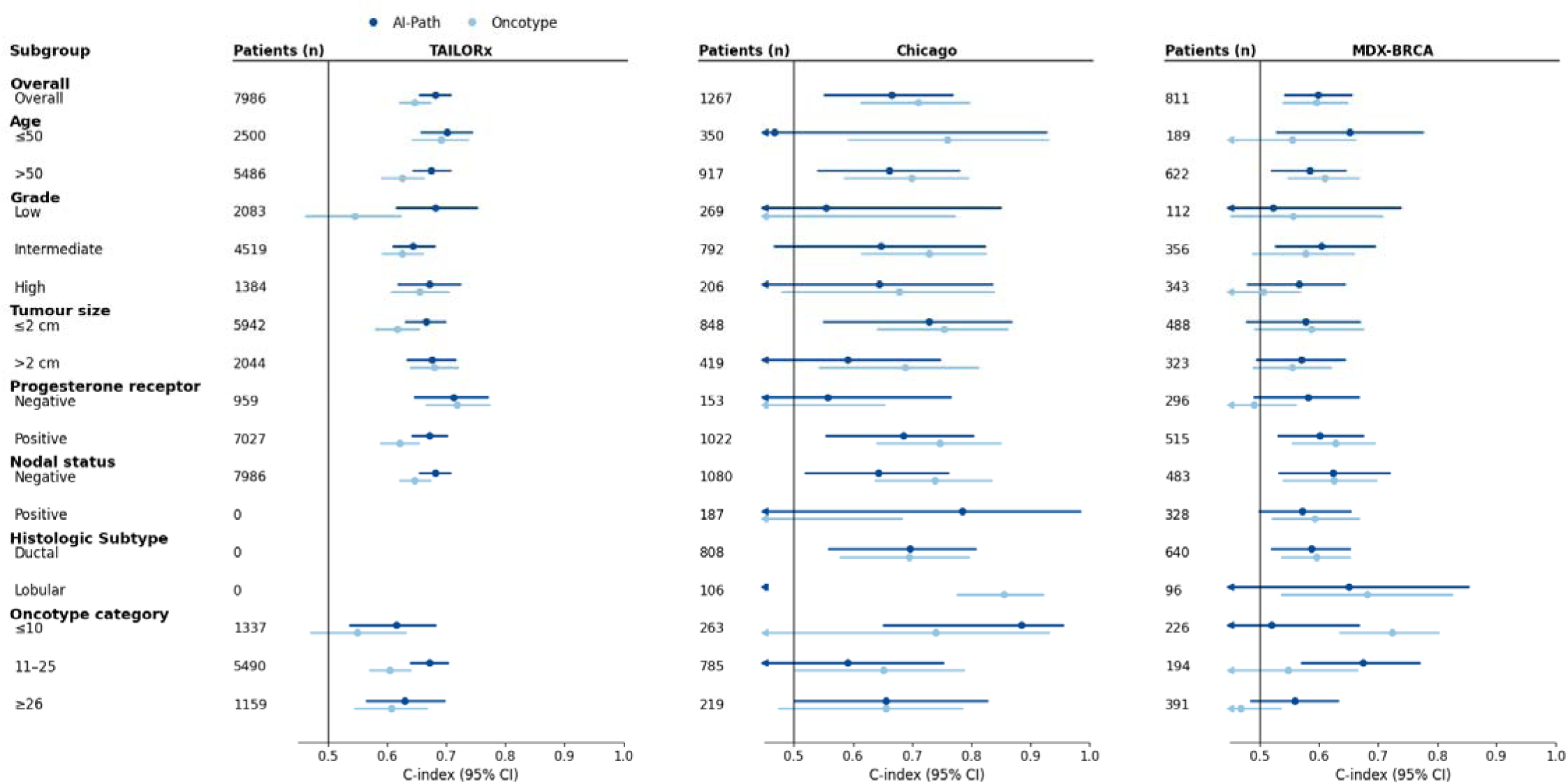
Subgroup Analysis of Prognostic Performance of AI-Path and Oncotype DX. Forest plots show the concordance index and 95% bootstrap confidence intervals for AI-Path and Oncotype DX within predefined clinicopathologic subgroups in the TAILORx, Chicago, and MDX-BRCA cohorts.

**Supplemental Figure 2.**
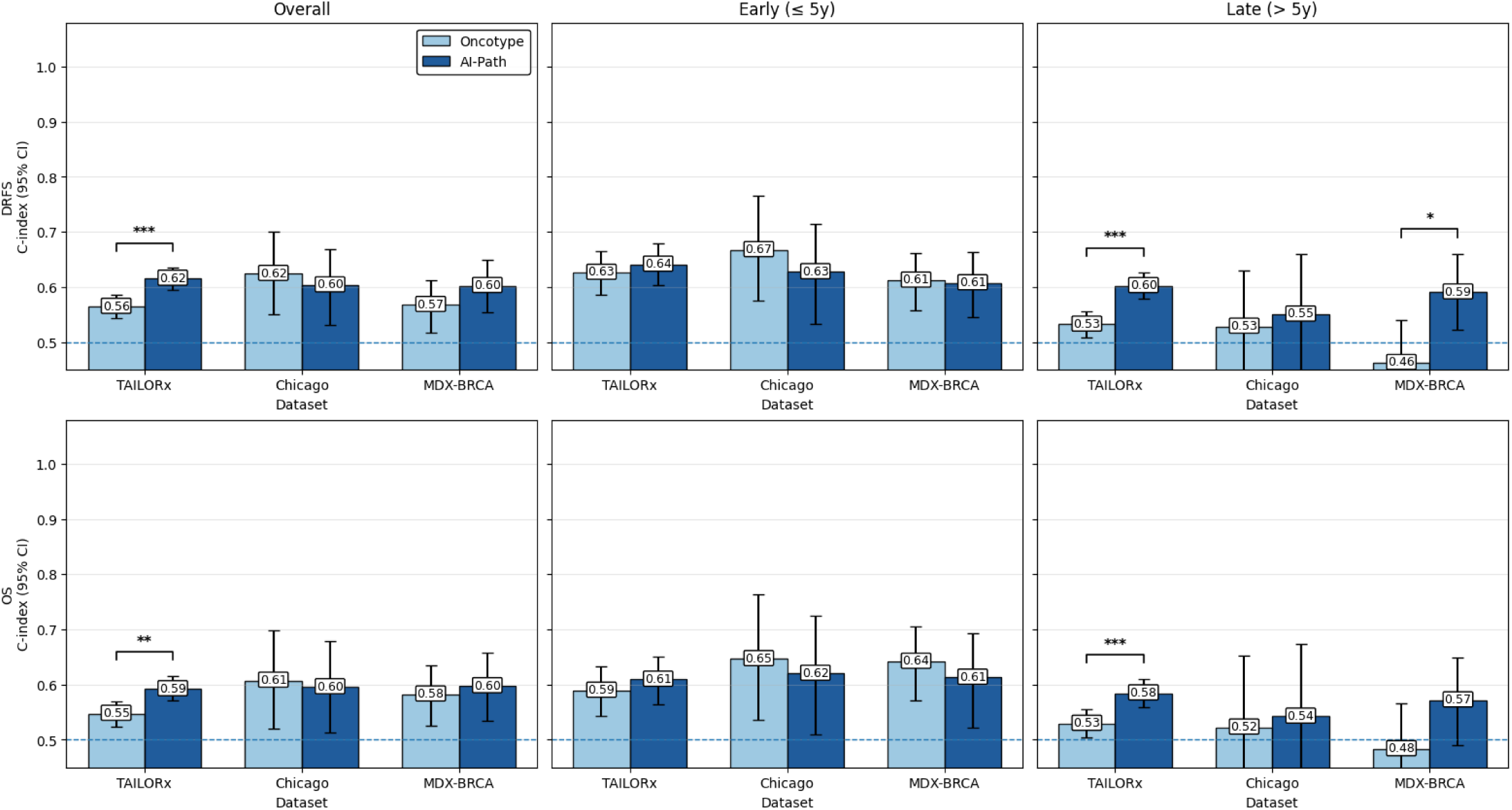
Prognostic Value of Oncotype and AI-Path for Distant Recurrence Free Survival and Overall Survival. The top row shows results for distant recurrence–free survival (DRFS), and the bottom row shows results for overall survival (OS). Within each endpoint, prognostic discrimination is shown for the overall cohort, events occurring within 5 years (early), and events occurring after 5 years (late). Bars indicate the estimated C-index for each model in each dataset, and error bars represent 95% bootstrap confidence intervals. Brackets and asterisks indicate the significance of the difference based on paired bootstrap testing within the same dataset and analysis window. * P < 0.05; ** P < 0.01; *** P < 0.001

**Supplemental Figure 3.**
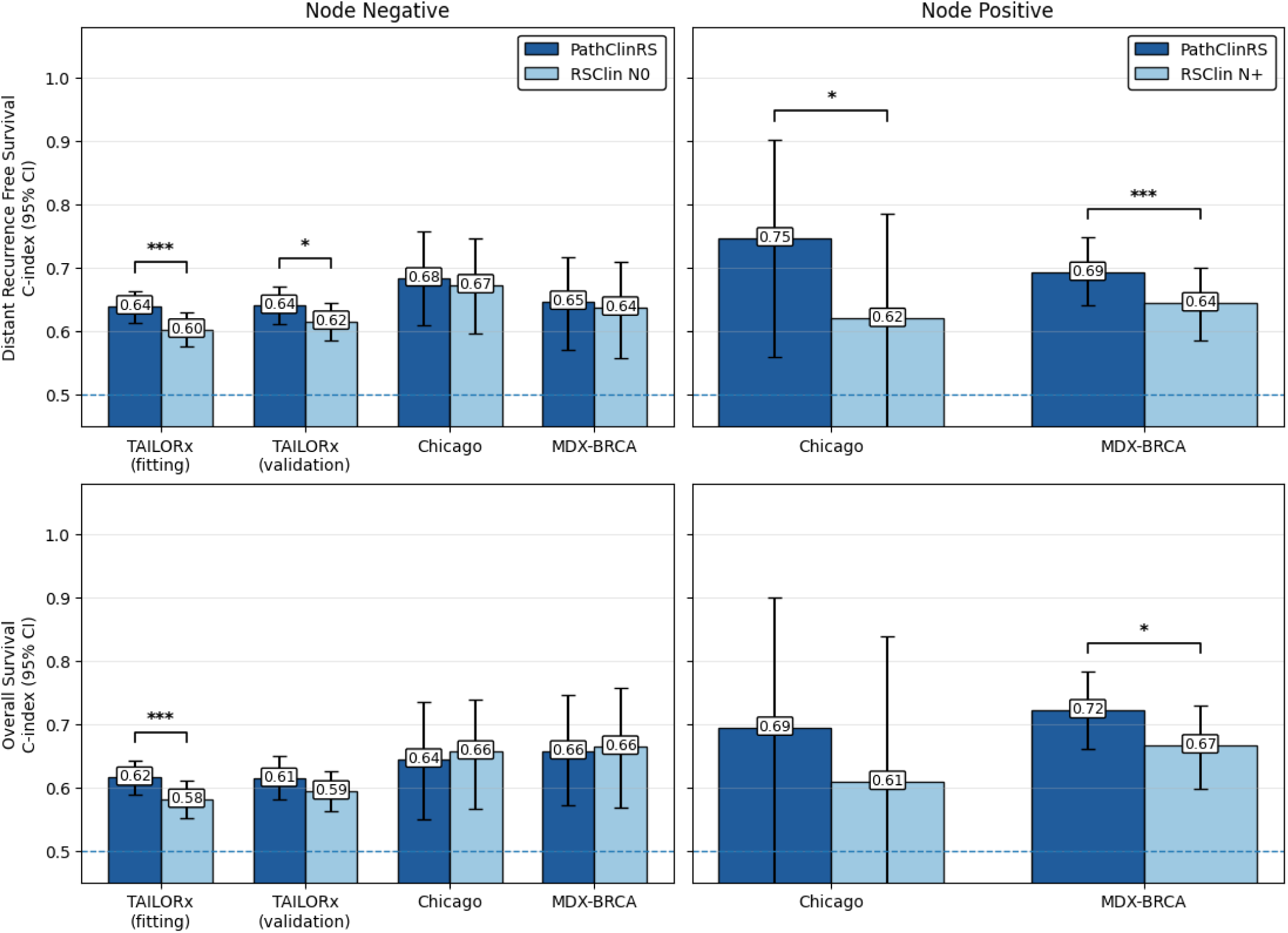
Prognostic Value of PathClinRS and RSClin for Distant Recurrence Free Survival and Overall Survival.

**Supplemental Figure 4.**
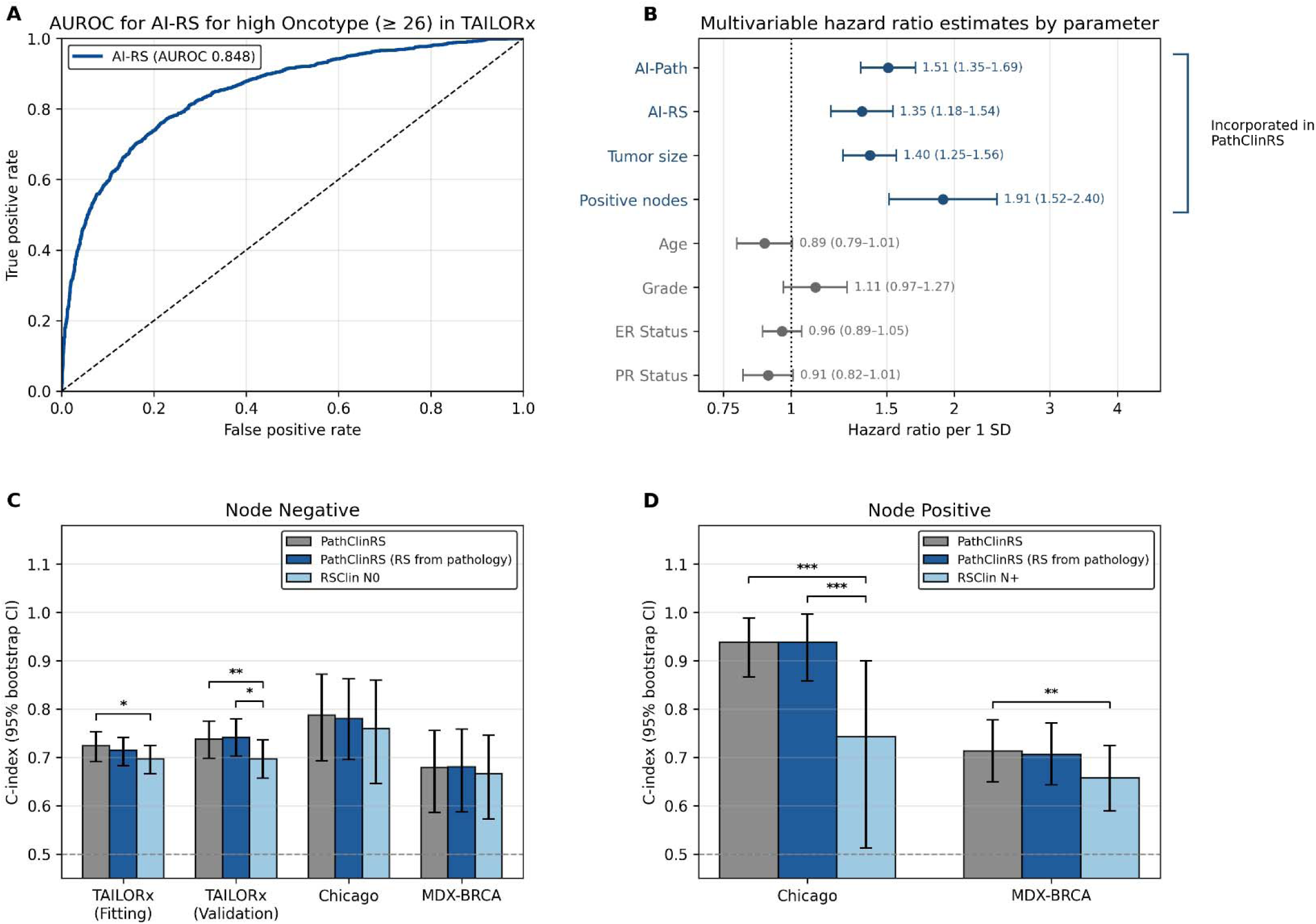
Evaluation of an AI Pathology Model for Oncotype Score for Genomic-Free Risk Prediction. (**A**) Receiver operating characteristic analysis showing the performance of AI-RS for predicting high-risk OncotypeDX score in TAILORx. (**B**) Multivariable Cox proportional hazards hazard-ratio estimates per 1 standard deviation demonstrate that the two computational pathology risk scores (AI-Path, trained to directly predict recurrence; and AI-RS, trained to predict Oncotype) are independently prognostic. (**C/D**) Predictive performance for distant recurrence measured by C-index in node-negative cohorts (**C**) and node-positive cohorts (**D**), comparing PathClinRS with Oncotype estimated from genomics versus pathology with RSClin N0 and RSClin N+. * P < 0.05; ** P < 0.01; *** P < 0.001

**Supplemental Figure 5.**
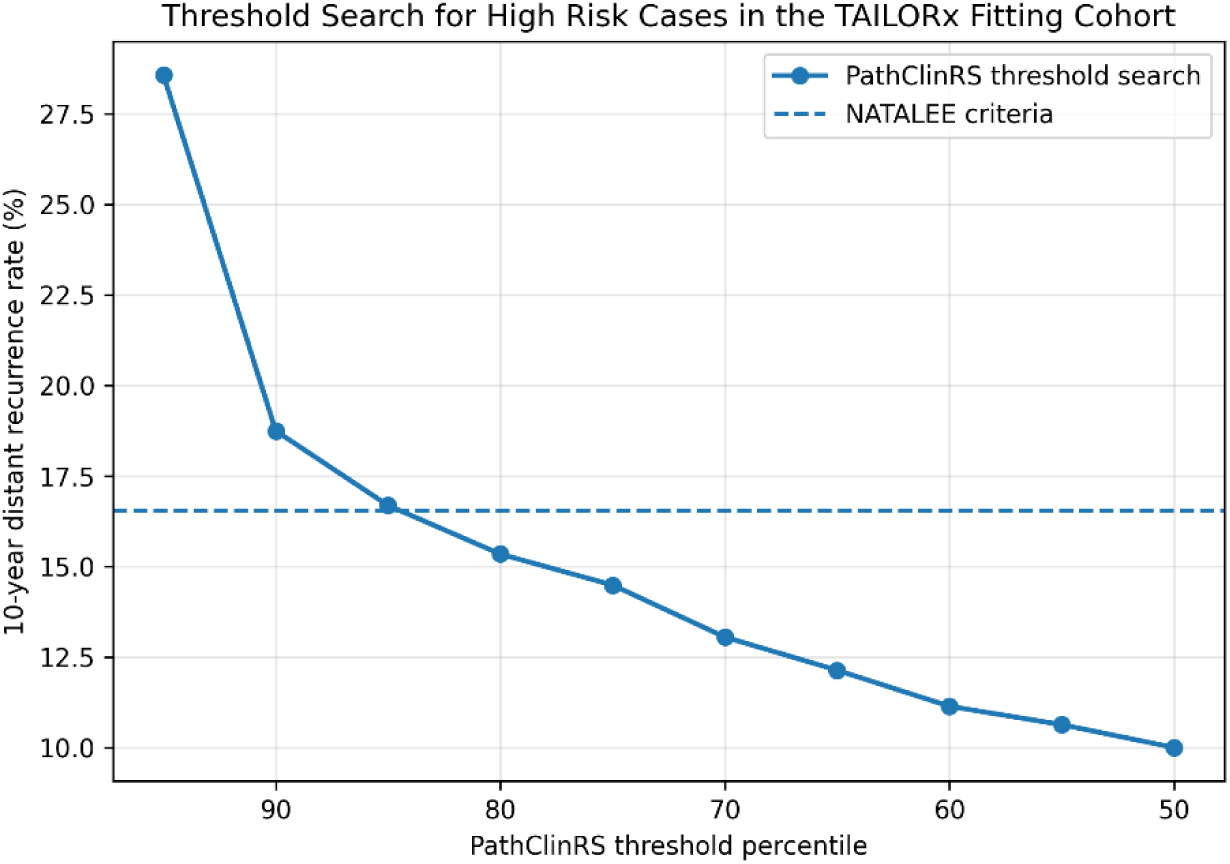
Threshold Search for 10-year Distant Recurrence in the TAILORx Model-Fitting Cohort. For each percentile cutoff of the PathClinRS score, the 10-year distant recurrence rate for cases above the threshold was estimated using Kaplan–Meier methods. The percentile cutoff identifying the largest group with a 10-year distant recurrence rate greater than that of NATALEE eligibility criteria was selected.

**Supplemental Figure 6.**
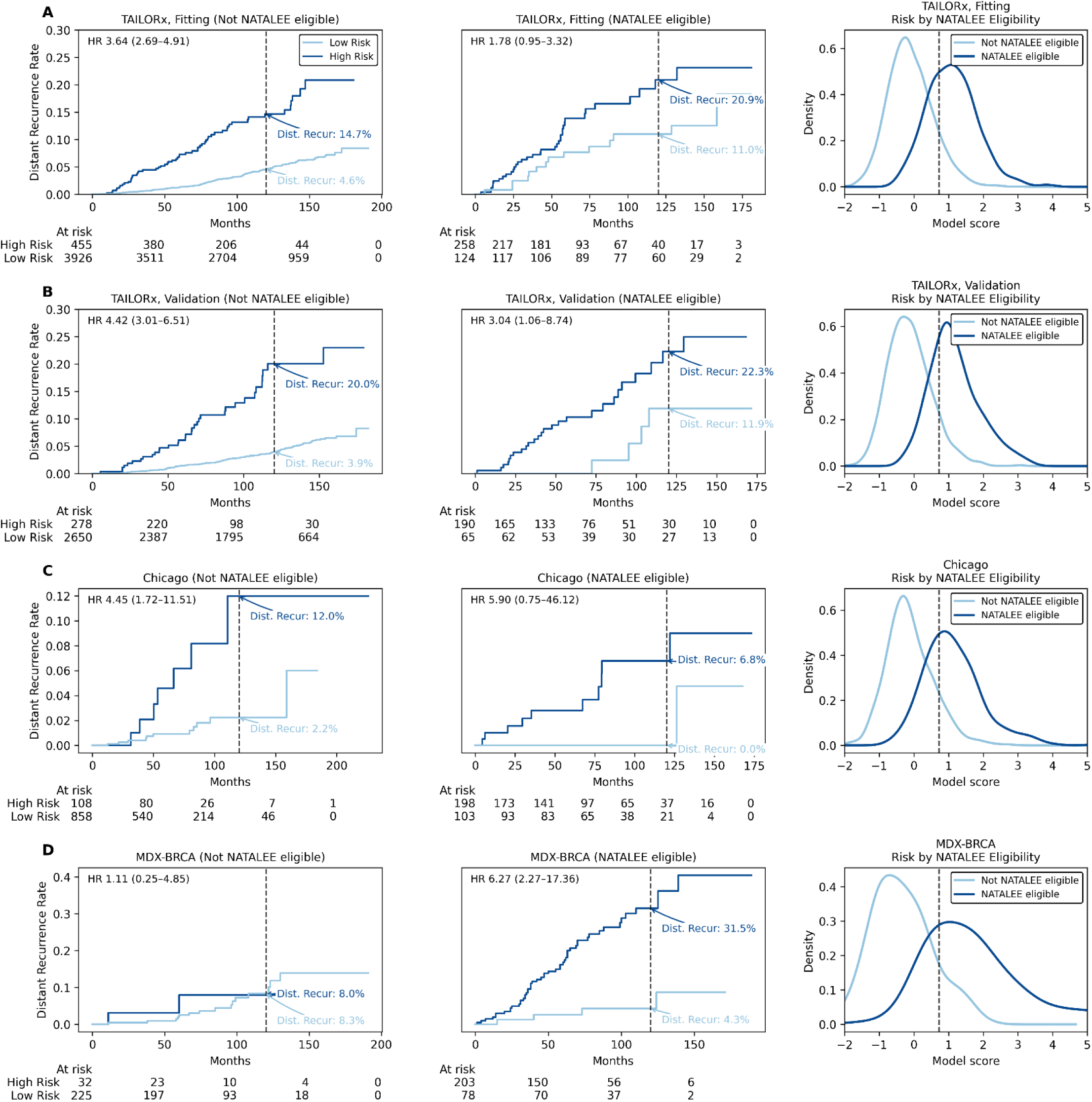
Prognostic stratification by PathClinRS within NATALEE-ineligible and NATALEE-eligible Subgroups. Kaplan–Meier estimates of distant recurrence are shown for the TAILORx model fitting cohort (**A**), TAILORx validation cohort (**B**), Chicago cohort (**C**), and MDX-BRCA cohort (**D**), stratified by both NATALEE eligibility (left / middle columns), and PathClinRS risk estimates (light / dark blue lines on graph).

**Supplemental Figure 7.**
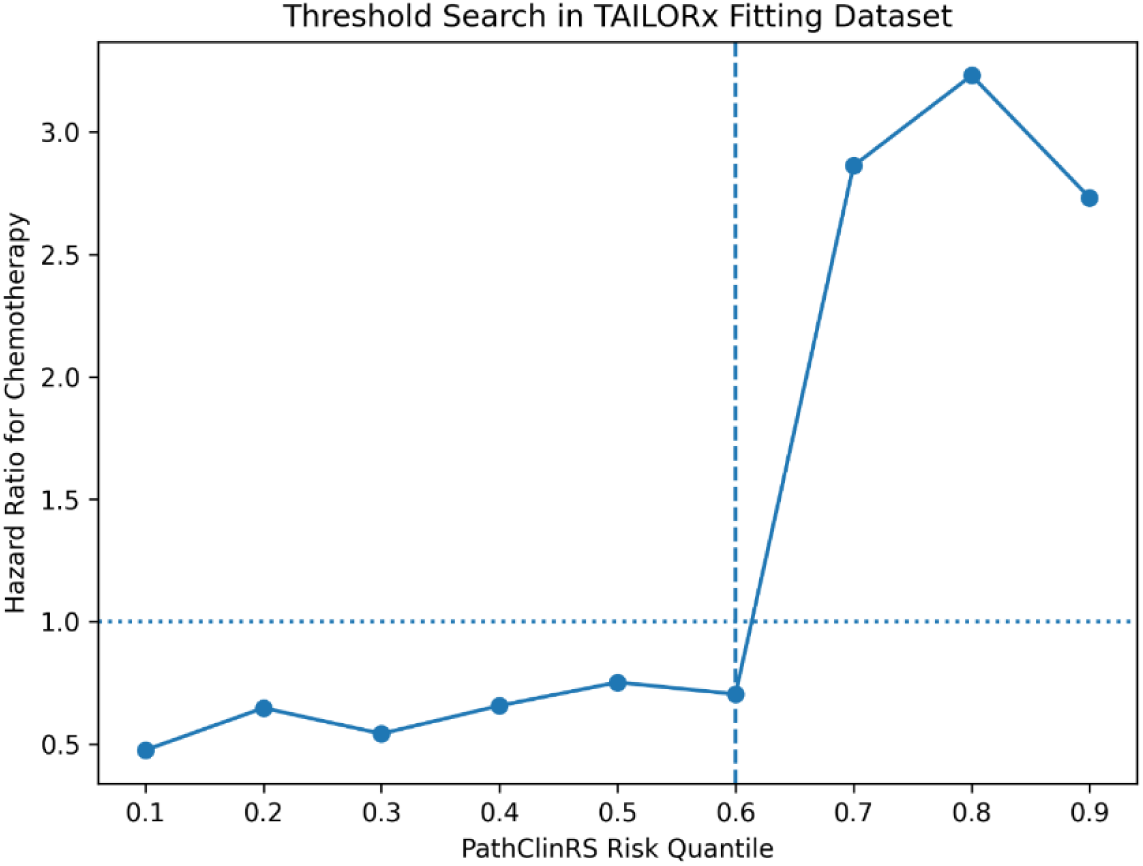
Threshold search for PathClinRS Chemotherapy Interaction. Among premenopausal patients in the TAILORx fitting cohort within arms B and C (randomized to chemoendocrine therapy or endocrine therapy alone), candidate PathClinRS cutoffs were evaluated across score quantiles (0.10 - 0.90). For a window around each candidate threshold (0.10 below to 0.10 above), the 10-year Kaplan-Meier distant recurrence risk was estimated separately by treatment arm within each risk group to determine the hazard ratio for chemotherapy benefit. The threshold which maximized the proportion of low risk patients while maintaining a lack of chemotherapy benefit was chosen.

**Supplemental Figure 8.**
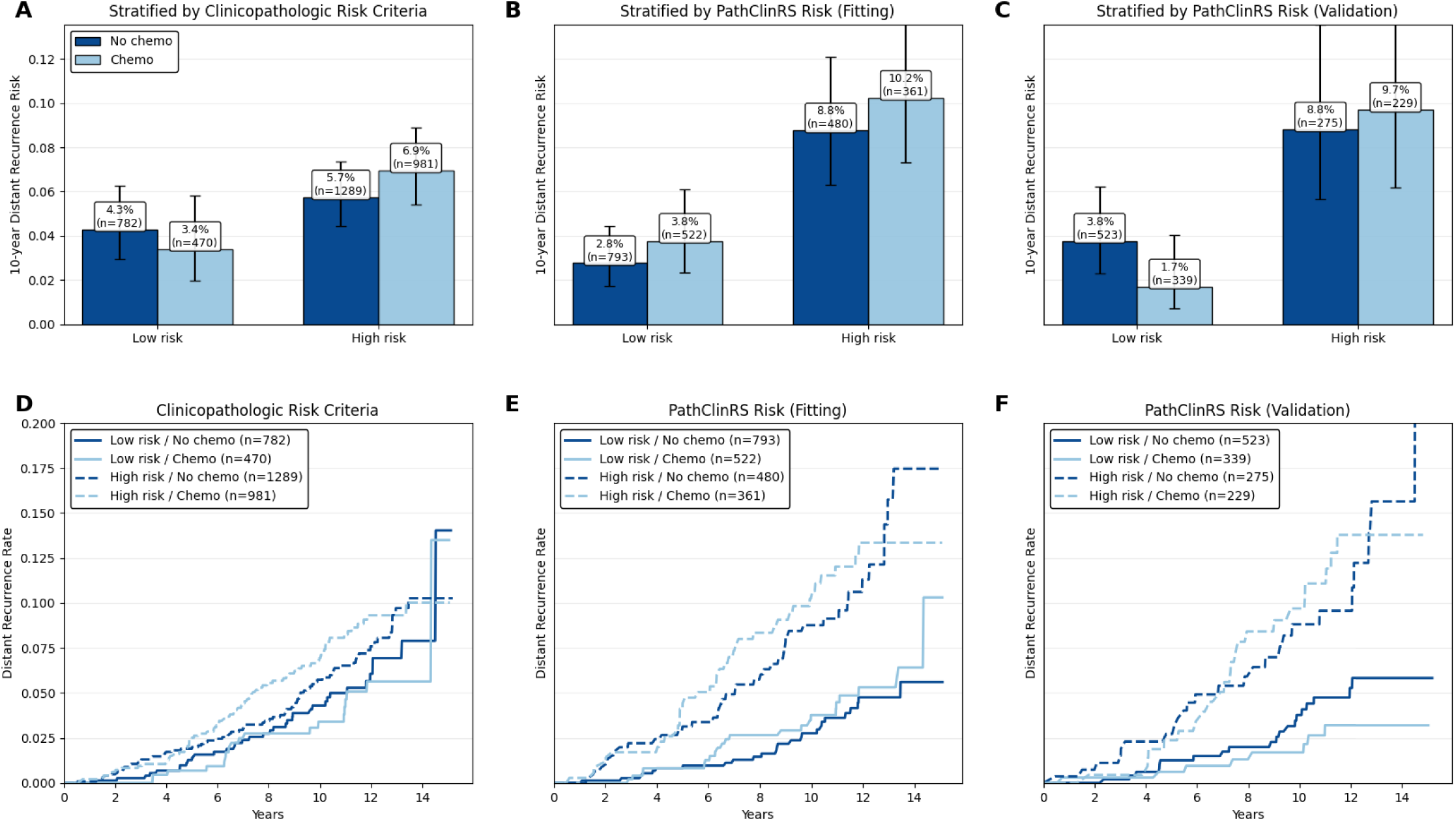
Lack of Chemotherapy Benefit in Intermediate Risk Postmenopausal Patients in TAILORx. (**A-C**) Bar plots showing Kaplan-Meier-estimated 10-year distant recurrence risk in low- and high-risk post-menopausal patients, further stratified by chemotherapy receipt. (**A**) Risk classification by clinicopathologic risk criteria, where high risk is defined as RS 21-25 or RS 16-20 with high clinical risk. (**B/C**) Risk classification by PathClinRS. (**D-F**) Kaplan-Meier curves for distant recurrence over time for the corresponding risk groups and chemotherapy strata for (**D**) clinicopathologic risk criteria and (**E**) PathClinRS risk group.

**Supplemental Table 1.** Cross-validated C-Index of Model Hyperparameters in the CALGB 9344 Training Set. C-index estimates for distant recurrence free interval are shown as raw averages at the best performing epoch. The best performing foundation model was first selected, and subsequently the learning rate for this model was optimized.

| Foundation Model | C-index |
| --- | --- |
| UNI | 0.613 |
| UNI2 | 0.615 |
| VIRCHOW2 | 0.605 |
| HOptimus0 | 0.617 |
| Learning Rate | C-index |
| 0.001 | 0.617 |
| 0.0005 | 0.621 |
| 0.0001 | 0.619 |
| 0.00005 | 0.616 |

**Supplemental Table 2.** C-index comparison of AI-Path and Oncotype by dataset and recurrence timeframe. C-index estimates for distant recurrence free interval are shown, and confidence intervals and p-values were determined through 1000x bootstrapping.

| Dataset | Timeframe | CI (95% CI) AI-Path | CI (95% CI) Oncotype | p-value |
| --- | --- | --- | --- | --- |
| TAILORx | Overall | 0.682 (0.655, 0.707) | 0.647 (0.620, 0.674) | 0.038 |
|  | Early | 0.708 (0.670, 0.745) | 0.728 (0.689, 0.764) | 0.382 |
|  | Late | 0.656 (0.623, 0.689) | 0.567 (0.529, 0.606) | <0.001 |
| Chicago | Overall | 0.665 (0.552, 0.768) | 0.710 (0.613, 0.796) | 0.492 |
|  | Early | 0.666 (0.501, 0.802) | 0.742 (0.626, 0.847) | 0.342 |
|  | Late | 0.663 (0.504, 0.815) | 0.631 (0.449, 0.787) | 0.758 |
| MDX-BRCA | Overall | 0.599 (0.541, 0.654) | 0.595 (0.539, 0.647) | 0.842 |
|  | Early | 0.600 (0.530, 0.673) | 0.641 (0.578, 0.699) | 0.358 |
|  | Late | 0.597 (0.515, 0.681) | 0.464 (0.382, 0.552) | <0.001 |

**Supplemental Table 3.**
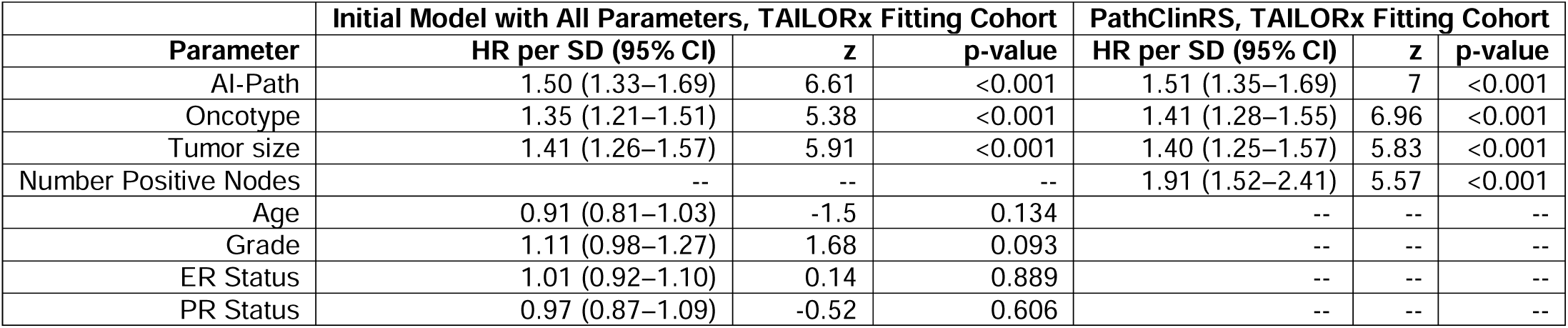
Hazard Ratios for Clinical, Pathologic, and Genomic Parameters in TAILORx. The initial model was fit with a comprehensive list of clinical, pathologic, and genomic parameters, with independently predictive parameters chosen for the final PathClinRS model. The hazard ratio for number of involved nodes was estimated from a two-parameter model incorporating tumor size and number of nodes using the data from CALGB 9344 and the TAILORx fitting cohort.

**Supplemental Table 4.** Overall Recurrence Discrimination in Node-Negative and Node-Positive Cohorts. For the N0 population, RSClin corresponds to the N0 model; for the N1+ population, RSClin corresponds to the N+ model.

| Population | Dataset | C-index (95% CI) PathClinRS | C-index (95% CI) RSCLin N0 / N+ | p-value |
| --- | --- | --- | --- | --- |
| N0 | TAILORx (Fitting) | 0.72 (0.69-0.75) | 0.70 (0.67-0.72) | 0.016 |
| N0 | TAILORx (Validation) | 0.74 (0.70-0.77) | 0.70 (0.66-0.74) | 0.004 |
| N0 | Chicago | 0.79 (0.69-0.87) | 0.76 (0.65-0.86) | 0.404 |
| N0 | MDX-BRCA | 0.68 (0.59-0.76) | 0.67 (0.57-0.75) | 0.502 |
| N1+ | Chicago | 0.94 (0.87-0.99) | 0.74 (0.51-0.90) | <0.001 |
| N1+ | MDX-BRCA | 0.71 (0.65-0.78) | 0.66 (0.59-0.72) | 0.004 |

**Supplemental Table 5.**
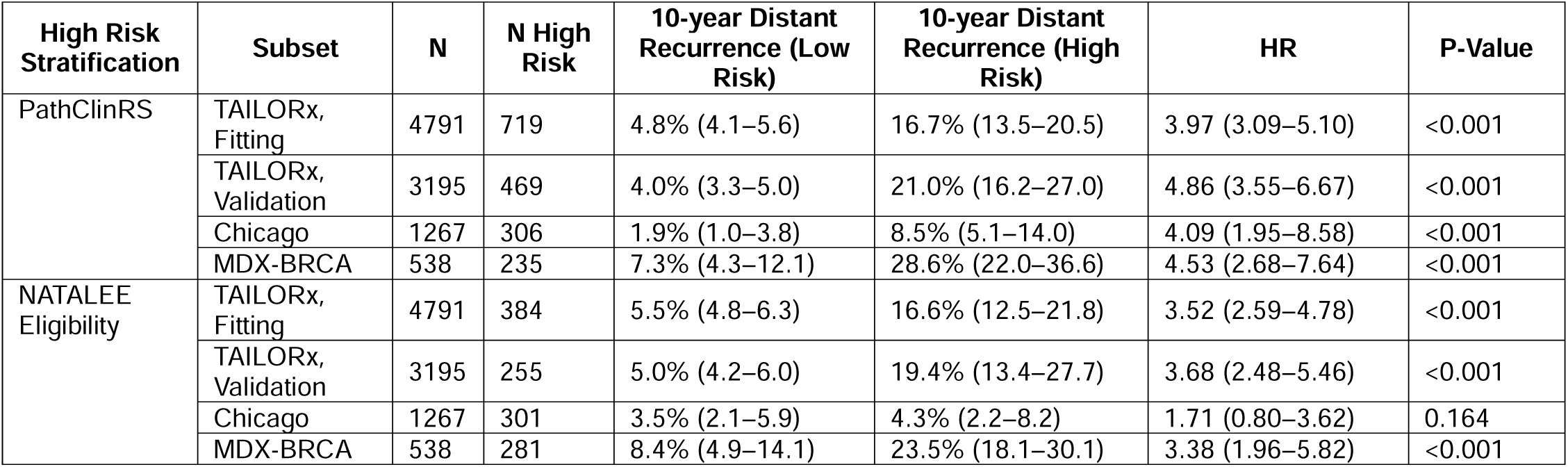
Distant Recurrence Estimates and Hazard Ratios for PathClinRS Risk Groups and NATALEE Trial Eligible / Ineligible Patients.

| High Risk Stratification | Subset | N | N High Risk | 10-year Distant Recurrence (Low Risk) | 10-year Distant Recurrence (High Risk) | HR | P-Value |
| --- | --- | --- | --- | --- | --- | --- | --- |
| PathClinRS | TAILORx, Fitting | 4791 | 719 | 4.8% (4.1–5.6) | 16.7% (13.5–20.5) | 3.97 (3.09–5.10) | <0.001 |
|  | TAILORx, Validation | 3195 | 469 | 4.0% (3.3–5.0) | 21.0% (16.2–27.0) | 4.86 (3.55–6.67) | <0.001 |
|  | Chicago | 1267 | 306 | 1.9% (1.0–3.8) | 8.5% (5.1–14.0) | 4.09 (1.95–8.58) | <0.001 |
|  | MDX-BRCA | 538 | 235 | 7.3% (4.3–12.1) | 28.6% (22.0–36.6) | 4.53 (2.68–7.64) | <0.001 |
| NATALEE Eligibility | TAILORx, Fitting | 4791 | 384 | 5.5% (4.8–6.3) | 16.6% (12.5–21.8) | 3.52 (2.59–4.78) | <0.001 |
|  | TAILORx, Validation | 3195 | 255 | 5.0% (4.2–6.0) | 19.4% (13.4–27.7) | 3.68 (2.48–5.46) | <0.001 |
|  | Chicago | 1267 | 301 | 3.5% (2.1–5.9) | 4.3% (2.2–8.2) | 1.71 (0.80–3.62) | 0.164 |
|  | MDX-BRCA | 538 | 281 | 8.4% (4.9–14.1) | 23.5% (18.1–30.1) | 3.38 (1.96–5.82) | <0.001 |

**Supplemental Table 6.** Distant Recurrence Risks and Hazard Ratios for NATALEE-Eligible and Ineligible Patients Stratified by PathClinRS Risk Group.

|  | Subset | N | N High Risk | 10-year Distant Recurrence (Low Risk) | 10-year Distant Recurrence (High Risk) | HR | P-Value |
| --- | --- | --- | --- | --- | --- | --- | --- |
| NATALEE Eligible | TAILORx, Fitting | 384 | 259 | 11.0% (6.3–18.6) | 20.9% (14.8–29.1) | 1.78 (0.95–3.32) | 0.072 |
|  | TAILORx, Validation | 255 | 190 | 11.9% (4.6–28.9) | 22.3% (14.8–32.8) | 3.04 (1.06–8.74) | 0.039 |
|  | Chicago | 301 | 198 | 0.0% (0.0–0.0) | 6.8% (3.5–13.0) | 5.90 (0.75–46.12) | 0.091 |
|  | MDX-BRCA | 281 | 203 | 4.3% (1.4–12.9) | 31.5% (24.2–40.3) | 6.27 (2.27–17.36) | <0.001 |
| NATALEE Ineligible | TAILORx, Fitting | 4407 | 460 | 4.6% (3.9–5.4) | 14.7% (11.2–19.1) | 3.64 (2.69–4.91) | <0.001 |
|  | TAILORx, Validation | 2940 | 279 | 3.9% (3.2–4.8) | 20.0% (14.2–27.8) | 4.42 (3.01–6.51) | <0.001 |
|  | Chicago | 966 | 108 | 2.2% (1.1–4.4) | 12.0% (5.3–26.0) | 4.45 (1.72–11.51) | 0.002 |
|  | MDX-BRCA | 257 | 32 | 8.3% (4.7–14.7) | 8.0% (2.0–29.2) | 1.11 (0.25–4.85) | 0.892 |

**Supplemental Table 7.** Distant Recurrence Risk and Chemotherapy Benefit for Premenopausal Patients Stratified by Clinicopathologic and PathClinRS Risk. Clinicopathologic risk criteria and PathClinRS-based risk groups are shown, with PathClinRS evaluated in the model fitting and validation cohorts. Hazard ratios were estimated from fully parameterized Cox proportional hazards interaction models including terms for chemotherapy, risk group, and their interaction; within-group chemotherapy hazard ratios were derived from the fitted model coefficients. Interaction P values correspond to the chemotherapy-by-risk-group interaction term.

| Criteria | Dataset | Risk group | Chemo | N | 10-year distant recurrence rate, % (95% CI) | Chemo HR (95% CI) | P value | Interaction P value |
| --- | --- | --- | --- | --- | --- | --- | --- | --- |
| Clinicopathologic Risk Criteria | TAILORx | Low Risk | No | 424 | 1.6 (0.7 - 3.5) | -- | -- |  |
|  |  |  | Yes | 288 | 2.2 (0.9 - 5.2) | 1.44 (0.50 - 4.10) | 0.497 |  |
|  |  | High risk | No | 634 | 8.8 (6.7 - 11.5) | -- | -- |  |
|  |  |  | Yes | 622 | 5.1 (3.5 - 7.5) | 0.58 (0.37 - 0.90) | 0.016 | 0.115 |
| PathClinRS Risk | Fitting | Low Risk | No | 431 | 2.2 (1.1 - 4.4) | -- | -- |  |
|  |  |  | Yes | 357 | 2.7 (1.4 - 5.3) | 1.46 (0.58 - 3.70) | 0.426 |  |
|  |  | High risk | No | 201 | 13.5 (9.2 - 19.5) | -- | -- |  |
|  |  |  | Yes | 168 | 6.6 (3.3 - 12.9) | 0.45 (0.22 - 0.93) | 0.031 | 0.051 |
|  | Validation | Low Risk | No | 305 | 2.4 (1.1 - 5.3) | -- | -- |  |
|  |  |  | Yes | 251 | 1.9 (0.7 - 5.1) | 0.90 (0.31 - 2.60) | 0.848 |  |
|  |  | High risk | No | 121 | 15.7 (9.9 - 24.6) | -- | -- |  |
|  |  |  | Yes | 134 | 9.7 (5.4 - 17.0) | 0.57 (0.27 - 1.22) | 0.145 | 0.489 |

**Supplemental Table 8.** Distant Recurrence Risk and Chemotherapy Benefit for Postmenopausal Patients Stratified by Clinicopathologic and PathClinRS Risk. Clinicopathologic risk criteria and PathClinRS-based risk groups are shown, with PathClinRS evaluated in the model fitting and validation cohorts. Hazard ratios were estimated from fully parameterized Cox proportional hazards interaction models including terms for chemotherapy, risk group, and their interaction; within-group chemotherapy hazard ratios were derived from the fitted model coefficients. Interaction P values correspond to the chemotherapy-by-risk-group interaction term.

| Criteria | Dataset | Risk group | Chemo | N | 10-year distant recurrence rate, % (95% CI) | Chemo HR (95% CI) | P value | Interaction P value |
| --- | --- | --- | --- | --- | --- | --- | --- | --- |
| Clinicopathologic Risk Criteria | TAILORx | Low Risk | No | 782 | 4.3 (2.9 - 6.3) | -- | -- |  |
|  |  |  | Yes | 470 | 3.4 (2.0 - 5.8) | 0.83 (0.47 - 1.44) | 0.5 |  |
|  |  | High risk | No | 1289 | 5.7 (4.5 - 7.4) | -- | -- |  |
|  |  |  | Yes | 981 | 6.9 (5.4 - 8.9) | 1.16 (0.84 - 1.60) | 0.36 | 0.298 |
| PathClinRS Risk | Fitting | Low Risk | No | 793 | 2.8 (1.7 - 4.4) | -- | -- |  |
|  |  |  | Yes | 522 | 3.8 (2.3 - 6.1) | 1.24 (0.70 - 2.19) | 0.459 |  |
|  |  | High risk | No | 480 | 8.8 (6.3 - 12.1) | -- | -- |  |
|  |  |  | Yes | 361 | 10.2 (7.3 - 14.2) | 1.09 (0.70 - 1.68) | 0.711 | 0.716 |
|  | Validation | Low Risk | No | 523 | 3.8 (2.3 - 6.2) | -- | -- |  |
|  |  |  | Yes | 339 | 1.7 (0.7 - 4.0) | 0.59 (0.26 - 1.33) | 0.202 |  |
|  |  | High risk | No | 275 | 8.8 (5.7 - 13.6) | -- | -- |  |
|  |  |  | Yes | 229 | 9.7 (6.2 - 15.0) | 1.05 (0.60 - 1.85) | 0.865 | 0.252 |

## Notes

### Author Declarations

IRB of University of Chicago gave ethical approval for this work

